# Usability, acceptability and cost of the SD BIOLINE Ov16 rapid diagnostic test for onchocerciasis surveillance in endemic communities in the middle belt of Ghana

**DOI:** 10.1101/2024.05.07.24306977

**Authors:** Kenneth Bentum Otabil, María-Gloria Basáñez, Ameyaa Elizabeth, Michael Oppong, Prince Mensah, Richmond Gyasi-Ampofo, Emmanuel John Bart-Plange, Theophilus Nti Babae, Lydia Datsa, Andrews Agyapong Boakye, Michael Tawiah Yeboah, Prince Nyarko, Prince Charles Kudzordzi, Anabel Acheampong, Edwina Twum Blay, Henk D.F.H. Schallig, Robert Colebunders

## Abstract

**Background:** Previous studies in the Bono Region (middle belt) of Ghana have reported persistent *Onchocerca volvulus* infection and associated morbidities after nearly three decades of ivermectin treatment. This study aimed to assess the usability, acceptability and cost of the Ov16 SD BIOLINE rapid diagnostic test (Ov16 RDT) in onchocerciasis surveillance activities in the middle belt of Ghana.

**Methodology:** A cross-sectional study was conducted in 6 endemic communities in the Tain District and Wenchi Municipality. A total of 254 individuals (54% females; median age (range)=31 (5–83) years), agreed to participate in Ov16 RDT (100%), skin-snip microscopy (37%) and nodule palpation (100%). Post-test interviews were conducted for all 94 participants tested by all three diagnostics. A cost analysis based on testing 400 people was performed.

**Principal findings:** Ov16 seroprevalence was 23.6% (60/254, 95%CI=18.8%–29.2%); microfilarial prevalence 11.7% (11/94, 95%CI = 6.7%–19.8%) and nodule prevalence 5.5% (14/254, 95%CI=3.3%–9.0%). The proportion of Ov16 seropositive females (43/136, 31.6%) was twice that of males (17/117, 14.5%). Among 5–9-year-olds, Ov16 seroprevalence was 11.1% (3/27), microfilarial prevalence 23.1% (3/13) and nodule prevalence 3.7% (1/27). For the 94 participants with all three tests, there was no association between the results of Ov16 RDT, skin-snip microscopy and/or nodule palpation. Most participants and technicians preferred Ov16 RDT because of being less painful and invasive, easier to use and faster. Had 400 participants been tested, the total cost per individual would be US$24 (Ov16 RDT) and US$74 (skin-snip microscopy).

**Conclusions:** Ov16 RDT is more acceptable and affordable (a third of the cost) compared to skin-snipping for surveillance activities in transmission hotspots in Ghana.

**Author summary:** Onchocerciasis (River blindness) is a neglected tropical disease targeted by the World Health Organization for elimination of transmission in 12 endemic countries by 2030. There is a need for field-friendly, acceptable and affordable tools to monitor progress towards elimination. In Ghana, the SD BIOLINE Ov16 rapid diagnostic test (Ov16 RDT) has been used in several epidemiological surveys, but its usability, acceptability and cost have not been assessed. We studied 6 endemic communities with persistent infection after nearly three decades of ivermectin treatment. The prevalence of seropositivity by Ov16 RDT was twice the prevalence of skin-snip microscopy positivity and four times the prevalence of nodule-palpation positivity. For the individuals tested by all three diagnostics, we found no agreement between the results of Ov16 RDT and skin-snip microscopy (and/or nodule palpation), likely owing to the long-term treatment in the study area. The Ov 16 RDT was acceptable to both study participants and technicians because it was less painful and invasive, and yielded results more quickly. The cost of skin-snip microscopy would be thrice that of Ov16 RDT when testing 400 individuals. Ov16 RDT is more acceptable and less costly than skin-snipping for surveillance activities in transmission hotspots in Ghana.

## Introduction

Onchocerciasis is a severely debilitating, vector-borne neglected tropical disease (NTD) caused by infection with the filarial nematode *Onchocerca volvulus* and transmitted among humans by the bites of *Simulium* blackflies. Although the great majority (99%) of the cases are found in sub-Saharan Africa (SSA), the infection also occurs in Yemen and in two (Venezuela and Brazil).out of the six original endemic countries in Latin America [1]. In 2017, it was estimated that at least 220 million people required preventive chemotherapy against onchocerciasis, and according to the Global Burden of Disease (GBD) Study for that year, 14.6 million of those infected had skin disease and 1.15 million had vision loss [2]. In 2019, the GBD Study estimated that 19.1 (95% Uncertainty Interval, UI = 17.2–20·9) million people were infected, and that the disease was responsible for 1.23 (95%UI = 0·765–1·82) million disability-adjusted life-years (DALYs) [3]. Preventive chemotherapy relies on mass drug administration (MDA) delivered, in Africa, as community-directed treatment with ivermectin (CDTI), predominantly annually, but in several foci of some countries (such as Ethiopia, Ghana, Nigeria, Togo and Uganda), also 6-monthly (biannually). Ivermectin is mainly microfilaricidal, and therefore CDTI programmes are typically prolonged, partly due to the long lifespan of adult *O. volvulus* [4]. Elimination (interruption) of transmission (EOT) has been reported for some foci of Mali, Senegal, Nigeria and Sudan, without vector control [5].

In consequence, the World Health Organization (WHO) 2021–2030 NTD roadmap has proposed that 12 endemic countries be verified for EOT of onchocerciasis by 2030 [6]. The WHO has delineated three distinct phases in onchocerciasis elimination programmes: I) the treatment phase with CDTI, during which monitoring and evaluation are conducted periodically to ascertain progress towards transmission suppression and ultimately interruption; II) the post-treatment surveillance (PTS) phase, during which CDTI is no longer delivered but indicators of exposure and transmission are evaluated to confirm that EOT has been reached, and III) the post-elimination surveillance (PES) phase, to alert of potential resurgence of transmission or re-introduction of infection [7]. These distinct phases require different diagnostic tools that allow transitioning from one to the next. During phase I, and until Stop-MDA surveys are conducted, the gold standard for parasitological diagnosis and calculation of epidemiological indicators (microfilarial prevalence and load), has been the detection and enumeration of *O. volvulus* microfilariae by the so-called skin-snip microscopy method [8]. Skin-snip microscopy is highly specific provided that microfilarial morphometric characteristics are assessed, and able to detect active infection (as microfilariae are the embryonic progeny of reproductively active adult worms), but its sensitivity (based on two snips) decreases as microfilarial prevalence and load declines due to CDTI [9]. The procedure is also mildly invasive and resource-intensive, and its acceptability diminishes in communities where decades of skin-snipping have occurred [10].

Skin-snip Polymerase Chain Reaction (PCR) has been suggested to circumvent the problem of low sensitivity posed by dwindling skin microfilarial loads, to confirm the infection status of Ov16-seropositive children, or when species-specific identification of microfilariae may be necessary [11]. However, it is not practical for large-scale community surveillance in SSA because it is expensive to purchase and operate and requires highly-skilled personnel. Serological, enzyme-linked immunosorbent assay (ELISA) tests based on the detection and quantification of IgG antibodies to the *O. volvulus* Ov16 recombinant antigen have shown, in longitudinal human and chimpanzee studies, that Ov16 seropositivity may precede skin-snip positivity, providing a marker of exposure to early infection stages [12]. Other studies, also in chimpanzees, have indicated that Ov16 (IgG4) seroconversion coincides with the onset of patency (detectable skin microfilariae) [13], but as its duration may exceed that of patent infection, Ov16 seropositivity may not yield a useful marker of active infection. Besides, ELISA-based serological tests also pose challenges including the need for laboratory analysis and the lack of a standardized, commercially-available Ov16 ELISA test, resulting in marked inter-laboratory variation [14]. A field-friendly, rapid diagnostic test (RDT) with the potential for integration into current onchocerciases surveillance activities in endemic countries, was developed and made commercially available (SD BIOLINE Onchocerciasis IgG4 rapid test) [14,15]. The performance of this Ov16 RDT has been evaluated in the field and current research priorities focus on operational and implementation research to demonstrate its usefulness, particularly in communities with low infection prevalence as a result of many years of CDTI [16].

In Ghana, ivermectin MDA has been implemented for more than 30 years, having started in the late 1980s [17]. In 2009, a nationwide rapid epidemiological mapping of onchocerciasis (REMO) evaluation was conducted to provide updated information on the distribution of the disease. Areas with an infection (palpable nodule) prevalence in adult male samples above 20%, were allocated to a biannual treatment frequency, also considering a buffer zone of 20 km around these CDTI priority areas [18]. Forty-four districts were classified as meso- and hyperendemic and have received biannual MDA since 2010. Forty-one districts, which were re-classified as of low endemicity and were already on MDA before the REMO exercise, continued to receive annual MDA. From 2015 to 2020, efforts were intensified to accelerate the shift from onchocerciasis control to EOT. An operational threshold of microfilarial prevalence <1% was adopted, the implementation unit changed from community to sub-district, and the geographical and (reported) therapeutic coverage increased substantially. In 2017, an impact assessment survey was carried out using Ov16 serology in children aged <10 years and skin-snip microscopy in adults aged ≥20 years [19]. Together with historical data collected by the Onchocerciasis Control Programme in West Africa (OCP), the impact assessment helped the national NTD Programme to formulate strategies to reach EOT. Delineation epidemiological surveys were conducted in 50 hypoendemic districts that had never received ivermectin (not having been previously prioritized for control but in need of re-assessment for elimination), and treatment strategies and management decisions were redefined [18]. Currently, 137 districts require or are under biannual CDTI [19].

Previous studies in the Bono region of Ghana, in which some communities had been under biannual CDTI since 2010, revealed that *O. volvulus* infection and associated cutaneous and ocular morbidities persisted despite 27 years of treatment [19,20], and that despite high levels of reported coverage [20], treatment adherence was relatively poor [21]. Although such communities may not be ready to move from the treatment phase to Stop-MDA surveys, or may require alternative interventions to accelerate progress [22] it is important to understand the role of different diagnostics for surveillance activities.

As Ghana transits from the treatment phase to the next stages of the EOT pathway in some endemic areas, and to evaluate progress towards suppression and interruption of transmission, the suitability of current diagnostic tools to accompany such transition needs to be ascertained. Whilst studies from other countries have investigated the acceptability, usability and cost of the Ov16 RDT [10], no such study has been conducted in Ghana. Therefore, this study aimed to assess the feasibility of integrating Ov16 RDT into onchocerciasis surveillance activities in Ghana based on diagnostic attributes, technician and community acceptability, usability and cost.

## Materials and methods

### Study area

The study was performed in six communities, namely, four communities in the Tain District (Abekwai 2, Abekwai 3, Attakrom and Kokomba) and two communities in the Wenchi Municipality (Blibor and Jonnykrom/Adamakrom), in the Bono Region of Ghana. Fig 1 shows the locations of the study villages. The communities were selected based on the following criteria informed by previous studies [19,20]: i) they had persistent endemicity for *O. volvulus* infection, albeit with relatively low prevalence; ii) they had received ivermectin MDA for at least 20 years and biannual CDTI for about 10, and iii) they were situated close to fast-flowing rivers (the Subin, the Tain, or their tributaries) where the blackfly vectors breed.

**Fig 1.**
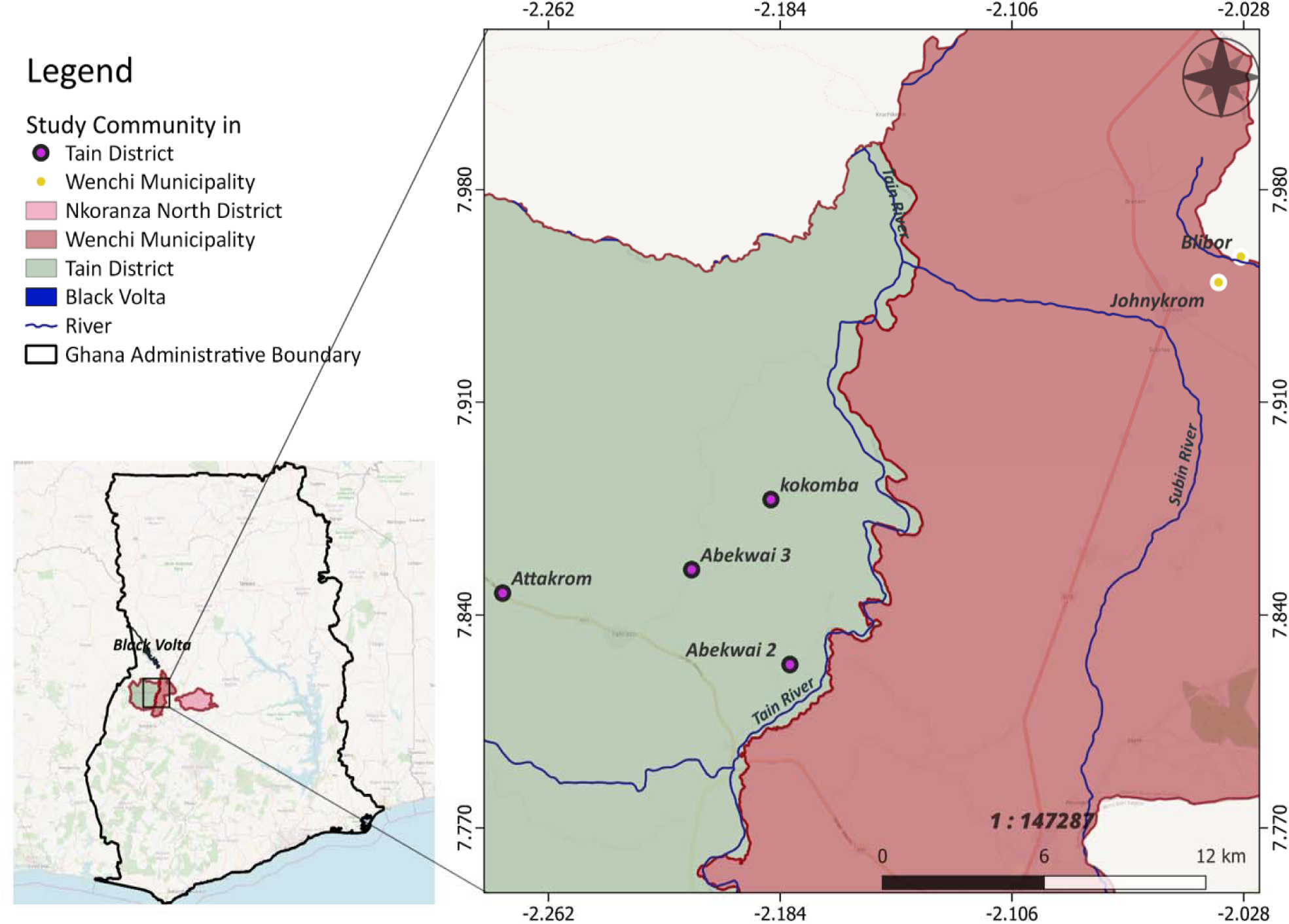
Map of study communities in the Tain District and Wenchi Municipality, Bono Region, Ghana. The light pink area to the right of the study area represents the Nkoranza North District.

### Study design and recruitment

This was a cross-sectional study, using both quantitative and qualitative approaches. Upon entry into the communities, the research team first met the chief and opinion leaders to explain the purpose, design, and procedures of the study. Once the community leaders agreed to participate and a date for the survey was decided, announcements were made via ‘gong-gong’ beaters and/or information centres to request community members to gather at a specific point in the village on the day. On the set day, the study was explained to the residents, and a total of 254 individuals aged ≥5 years agreed to participate in some or all components of the study, namely, a sociodemographic and health history questionnaire, Ov16 RDT testing (SD BIOLINE Onchocerciasis IgG4 Rapid Test), two skin-snip biopsies for microscopy, nodule palpation, and a post-test (exit) interview to assess their perceptions on the tests deployed and to understand in which tests they would be willing to participate in future surveys. Participants (or their parents/guardians) were asked to sign an informed consent form. Table 1 provides details on the numbers recruited at each community, their sex composition and median age, and the numbers examined for each onchocerciasis diagnostic. The study was conducted between June and August 2021.

**Table 1.** Characteristics of the population sampled for each study community in the Bono region of Ghana.

| Community | Recruited/<br>Population | Sex<sup>†</sup> |  | Median<br>age<sup>‡</sup><br>(range)<br>(Years) | No. Tested |  |  |
| --- | --- | --- | --- | --- | --- | --- | --- |
|  |  | Females | Males |  | Ov16<br>RDT<br>(%) | Skin snip<br>(%) | Nodules<br>(%) |
|  | (%) | (%) | (%) |  |  |  |  |
| Tain District |  |  |  |  |  |  |  |
| Abekwai 2 | 30/150<br>(20.0%) | 17<br>(56.7%) | 13<br>(43.3%) | 22<br>(5–61) | 30<br>(100%) | 11<br>(36.7%) | 30<br>(100%) |
| Abekwai 3 | 60/700<br>(8.6%) | 29<br>(48.3%) | 31<br>(51.7%) | 30<br>(5–83) | 60<br>(100%) | 23<br>(38.3%) | 60<br>(100%) |
| Attakrom | 41/800<br>(5.1%) | 21<br>(52.5%) | 19<sup>†</sup><br>(47.5%) | 47<br>(9–80) | 41<br>(100%) | 15<br>(36.6%) | 41<br>(100%) |
| Kokomba | 40/120<br>(33.3%) | 24<br>(60.0%) | 16<br>(40.0%) | 25<br>(7–60) | 40<br>(100%) | 16<br>(40.0%) | 40<br>(100%) |
| Wenchi Municipality |  |  |  |  |  |  |  |
| Blibor | 47/240<br>(19.6%) | 18<br>(38.3%) | 29<br>(61.7%) | 27<br>(7–80) | 47<br>(100%) | 12<br>(25.5%) | 47<br>(100%) |
| Johnykrom/<br>Adamakrom | 36/170<br>(21.2%) | 27<br>(75.0%) | 9<br>(25.0%) | 41<br>(6–72) | 36<br>(100%) | 17<br>(47.2%) | 36<br>(100%) |
| Total |  |  |  |  |  |  |  |
|  | 254/2,180<br>(11.7%) | 136<br>(53.8%) | 117<br>(46.3%) | 31<br>(5–83) | 254<br>(100%) | 94<br>(37.0%) | 254<br>(100%) |

<sup>†</sup>Of the 254 participants, sex was recorded for 253 (sex missing for one participant in Attakrom).
<sup>‡</sup>Age was recorded for 242 participants (age missing for one participant in each of Abekwai 2 and Abekwai 3, four participants in Attakrom, three in Kokomba and three in Johnykrom).

### Ethical considerations

Ethical clearance for this study was obtained from the Committee for Human Research and Ethics of the University of Energy and Natural Resources in Sunyani, Ghana, West Africa (approval number: CHRE/AP/012/021). The study was explained to the parents/guardians and school-age children in the local dialect (*Twi*). Informed written consent for children less than 18 years of age was given by their parents/guardians. Children gave oral assent to confirm their willingness to participate in the study. Participants were informed that they had the option of withdrawing at any stage of the study, without having to give any reasons, and without any consequences following withdrawal.

### Sociodemographic questionnaire

Participants were asked to state their sex and age, and respond to questions regarding whether they had been born in the village (yes or no); their duration of residence in the community (<1 year or ≥1 year); whether they lived close to a stream (yes or no), visited a stream (yes or no) and how often (every day or less than everyday), and their occupation (farming or non-farming), with the aim to obtain information indicative of exposure to explore its association with Ov16 status (positive or negative).

### Ov16 rapid diagnostic test

The deployment of the IgG4-based SD BIOLINE rapid diagnostic test (Ov16 RDT) followed the manufacturer’s instructions (Standard Diagnostics, Yongin-si, Gyeonggi-do, South Korea) and quality-assurance procedures. Briefly, the test cassette was labelled with the participant’s unique identification number. A finger of the participant was selected, cleaned with an alcohol swab and pricked with the lancet provided with the kit. The blood was then collected using a capillary tube and the blood was placed in the specimen well of the cassette. Four drops of assay diluent were also placed in the assay diluent well. RDT results were read after 20 minutes [10]. The product insert for the Ov16 RDT states that, compared to skin-snip microscopy as the gold standard, its sensitivity is 81.1% (95%CI = 70.7%–88.4%) and its specificity is 99.0% (95%CI = 94.8%–99.8%) using whole blood [10].

### Skin-snip microscopy

Skin snipping was performed as described in our previous studies [19,20]. Briefly, the left and right iliac crests of each participant were disinfected using alcohol swabs. Holth corneoscleral punches were used to take about 2mg of bloodless skin snips each from the left and right iliac crests. The snips were placed separately in microtitre wells containing 100 µl normal saline (NaCl 0.85%) and incubated at room temperature for 24 hours. After this period, the incubation medium was pipetted onto a microscope slide and examined using, firstly, a x10 and subsequently a x40 objective lens. If microfilariae (mf) had emerged, the participant was recorded as skin-snip positive.

### Nodule palpation

All participants were examined for palpable nodules by trained members of the medical team (including a doctor, nurse and laboratory technicians) with experience in detecting onchocercal nodules (onchocercomas). Palpations were done on bony protuberances of the ribs, iliac crests, sacrum and upper leg, but not at locations (neck, axillae, inguinal) typical for lymph nodes. Nodules were defined as ‘suspected’ onchocercomas (i.e., not confirmed by dissection or biopsy) which were firm, often flattened or bean-shaped, usually movable, non-tender and up to several centimetres in diameter.

### Usability and acceptability of Ov16 RDT

To assess the usability and acceptability of the RDT, all of the RDT technicians (n=9) who also had experience in skin-snip microscopy, and community residents (n=94) who participated in all three tests (RDT, skin-snip microscopy and nodule palpation) were interviewed (S1 Text). The interview guide was developed along the themes of preference and reasons for preference by participants and technicians regarding the use of Ov16 RDT for surveillance following [10]. The interviews were recorded using an audio recorder on Android phones and transcribed *verbatim*. Interview data were coded using content analysis based on key themes from the semi-structured interviews as done in [10].

### Economic costs

To evaluate the cost associated with the implementation of onchocerciasis surveillance programmes in Ghana using Ov16 RDT and skin-snip microscopy, a costing analysis was done based on 400 participants as the basis for our calculations to enable comparison of costs for each test without the effect of the numbers actually tested in any given survey. Briefly, the value of volunteers’ time as well as the surveillance costs were computed using the ‘ingredient’ approach, multiplying the input prices by the number of inputs used [23]. The major input prices for this analysis were the cost of each: a) Ov16 RDT (US$2.82 per cassette); b) Holth corneoscleral punch (US$423.5 per punch), and c) optical microscope (US$1,694). In addition, costs were captured for all activities conducted during the surveillance activity, including training, fieldwork and data reporting. Fieldwork cost categories were further split into transport and lodging, supplies, devices and instruments, and labour. An exchange rate of GHS7.08 (cedis) per US$ was used in agreement with the rate for 2021.

### Data analysis

Data were entered in purposely designed MS Excel spreadsheets (one for the epidemiology results and another for the cost menus). Analyses were conducted using Jamovi Desktop (version 2.3.19) and GraphPad Prism 8 for macOS (version 8.2.1). Two x two contingency tables for the (dichotomous) responses to the sociodemographic questionnaire and Ov16 status (positive or negative) were analysed using (two-tailed) Chi-squared test (without Yates continuity correction as all frequencies were greater than 1) [24]. Two x two tables for concordance were analysed for those participants who underwent all three tests, firstly comparing Ov16 with skin-snip results and secondly, comparing Ov16 with skin-snip and/or nodule-palpation results combined as ‘Oncho’ status. For such analyses we report the values of the following parameters: i) kappa statistic and its 95% confidence interval (95%CI), a measure of agreement between the results of two alternative techniques of categorical assessment that takes into account whether the agreement is occurring by chance; ii) the overall proportion of agreement (*p*_0_); iii) the proportion of positive agreement (*p*_pos_); iv) the proportion of negative agreement (*p*_neg_), v) the McNemar’s Chi-squared statistic with its P-value; and vi) the sensitivity, specificity, positive predictive value (PPV) and negative predictive value (NPV) [25,26]. When reporting proportions, 95%CIs were calculated using the Wilson score method [27]. Proportions were compared by assessing their 95%CIs or by using the z-statistic where appropriate (non-paired data), in which case asymptotic 95%CIs are reported [24]. The significance level for all analyses was set at 0.05. A sample size was not calculated a priori for the study sites; our sample is a convenience sample with an average sample size of 12% (range=5%–33%) of the community populations (Table 1). The number of participants recruited per village (at least 30 participants per community) is consistent with the usual number of individuals per village recruited for onchocerciasis epidemiological studies [28].

## Results

Of the total of 254 individuals, there were 253 with recorded sex, consisting of 136 (53.8%) females and 117 (46.3%) males. Age was recorded for 242 individuals, with a median age of 31 (range=5–83) years (Table 1). Respectively, 254 (100%), 94 (37%) and 254 (100%) individuals agreed to participate in the Ov16 RDT, skin-snip microscopy and nodule palpation tests. Therefore 94 participants underwent all three tests. The proportion of the population for whom microfilaridermia was assessed ranged from 26% to 47%.

Of the 254 individuals tested for Ov16 RDT, 60 (23.6%, 95%CI = 18.8 %–29.2%) were seropositive, compared to 11 (11.7%, 95%CI = 6.7%–19.8%) microfilaria-positive individuals out of the 94 with skin-snip results (z-value = 2.5; P-value = 0.0143). For palpable nodules, 14 out of 254 individuals examined were positive (5.5%, 95%CI = 3.3%–9.0%). The prevalence of palpable nodules was only marginally significantly different from that of microfilaridermia (z-value = 2, P-value = 0.0471), but highly significantly different from that of Ov16 (z-value = 5.8, P-value <0.0001).

The distribution of Ov16 status based on (dichotomised) sociodemographic information and infection status is presented in Table 2. The following variables were statistically significantly associated with Ov16 status (positive or negative): sex (female or male), age (<30 or ≥30 years), occupation (farming or not farming) and palpable nodules (positive or negative). Microfilaridermia and Ov16 statuses were not associated. Among seropositives, the proportion of females (71.7%, 95%CI = 59.2%–81.5%) was statistically significantly higher than that of males (28.3%, 95%CI = 18.5%–40.8%); the proportion of those aged ≥30 years (76.4%, 95%CI = 63.7%–85.6%) was higher than those aged <30 (23.6%, 95%CI = 14.4%–36.4%), and the proportion of farmers (84.8%, 95%CI = 73.5%–91.8%) was higher than that of non-farmers (15.3%, 95%CI = 8.2%–26.5%). The proportion of those without microfilaridermia (89.5%, 95%CI = 68.6%–97.1%) was higher than of those with a positive skin snip (10.5%, 95%CI = 2.9%–31.4%), and the proportion of those without palpable nodules (88.3%, 95%CI = 77.8%– 94.2%) was higher than of those presenting onchocercomas (11.7%, 95%CI = 5.8%–22.2%).

**Table 2.** Sociodemographic information, infection status and Ov16 RDT status of participants.

| Description [No. participants for whom data were collected] | No. Ov16 Negative (%) | No. Ov16 Positive (%) | $\chi^2$ statistic (P-value<sup>†</sup>) |
| --- | --- | --- | --- |
| Sex [253] |  |  |  |
| Female | 93 (36.8%) | 43 (17.0%) | 10.15<br>(0.0014) |
| Male | 100 (39.5%) | 17 (6.7%) |  |
| Age [242] |  |  |  |
| <30 years | 101 (41.7%) | 13 (5.4%) | 15.74<br>(0.0001) |
| ≥30 years | 86 (35.5%) | 42 (17.4%) |  |
| Duration of stay in the village [244] |  |  |  |
| Born in village and <1 year | 92 (37.7%) | 23 (9.4%) | 0.57<br>(0.4490) |
| ≥1 year | 98 (40.2%) | 31 (12.7%) |  |
| Proximity to stream [254] |  |  |  |
| Yes | 166 (65.4%) | 49 (19.3%) | 0.54<br>(0.4639) |
| No | 28 (11.0%) | 11 (4.3%) |  |
| Visit stream [236] |  |  |  |
| Yes | 133 (56.4%) | 40 (16.9%) | 0.13<br>(0.7162) |
| No | 47 (19.9%) | 16 (6.8%) |  |
| Frequency visiting stream [170] |  |  |  |
| Everyday | 47 (27.6%) | 9 (5.3%) | 1.04<br>(0.3073) |
| <Everyday | 88 (51.8%) | 26 (15.3%) |  |
| Occupation [251] |  |  |  |
| Farming | 127 (50.6%) | 50 (19.9%) | 7.51<br>(0.0061) |
| Non-farming | 65 (25.9%) | 9 (3.6%) |  |
| Presence of skin microfilariae [94] |  |  |  |
| Yes | 9 (9.6%) | 2 (2.1%) | 0.03<br>(0.8583) |
| No | 66 (70.2%) | 17 (18.1%) |  |
| Presence of palpable nodules [254] |  |  |  |
| Yes | 7 (2.8%) | 7 (2.8%) | 5.71<br>(0.0168) |
| No | 187 (73.6%) | 53 (20.9%) |  |

<sup>†</sup>P-value for the 2-tailed Chi-squared statistic for 2 x 2 contingency tables.

Also among the Ov16 seropositives, the proportion of those living close to streams (81,7%, 95%CI = 70.1%–89.4%) was higher than that of those who did not live close (18.3%, 95%CI = 10.6%–29.9%), and the proportion of those visiting streams (71.4%, 95%CI = 58.5%–81.6%) was higher than that of those who did not visit streams (28.6%, 95%CI =18.4%–41.5%). However, the proportion of those visiting streams daily was lower (25.7%, 95%CI = 14.2%– 42.1%) than of those visiting less frequently (74.3%, 95%CI = 57.9%–85.8%).

The two participants who were both Ov16 and skin-snip positive were females who had taken ivermectin in the previous year (2019). Of the 17 participants who were Ov16 positive but skin-snip negative, 13 (76.5%) were female and all 17 had taken ivermectin in the previous year.

### Prevalence by age and sex of test positivity for Ov16 RDT, skin-snip microscopy and nodule palpation

Table 3 presents the results of the age and sex distribution of Ov16 RDT, skin-snip and palpable nodule positivity. The prevalence of the three indicators tended to be higher among those aged ≥30 years, but confidence intervals were wide. The seroprevalence in this age group was 32.8% (95%CI = 25.3%–41.3%). In the 5–9 age group, 3/27 children (11.1%, 95%CI = 3.9%–28.1%) were Ov16 positive, 3/13 children (23.1%, 95%CI = 8.2%–50.3%) skin-snip positive, and 1/27 (3.7%, 95%CI = 0.7%–18.3%) nodule positive. The results by year of age for Ov16 were: 0/4 for 5-year-olds, 0/5 for 6-year-olds, 1 (boy)/9 for 7-year-olds, 0/3 for 8-year-olds and 2 (one boy and one girl)/6 for 9-year-olds. For skin-snip positivity, the results were: 0/3 for 5-year olds, 1 (boy)/4 for 6-year olds, 2 (one boy and one girl)/3 for 7-year olds, 0/2 for 8-year olds and 0/1 and 9-year olds. The nodule-positive child (boy) was aged 5. The prevalence of Ov16 seropositivity among females (31.6%) was twice as high as that among males (14.5%), and this difference was statistically significant (z-value = 3.2, P-value = 0.0014). Fig 2 presents Ov16 seropositivity by age group and sex for the 242 participants with recorded age.

**Fig 2.**
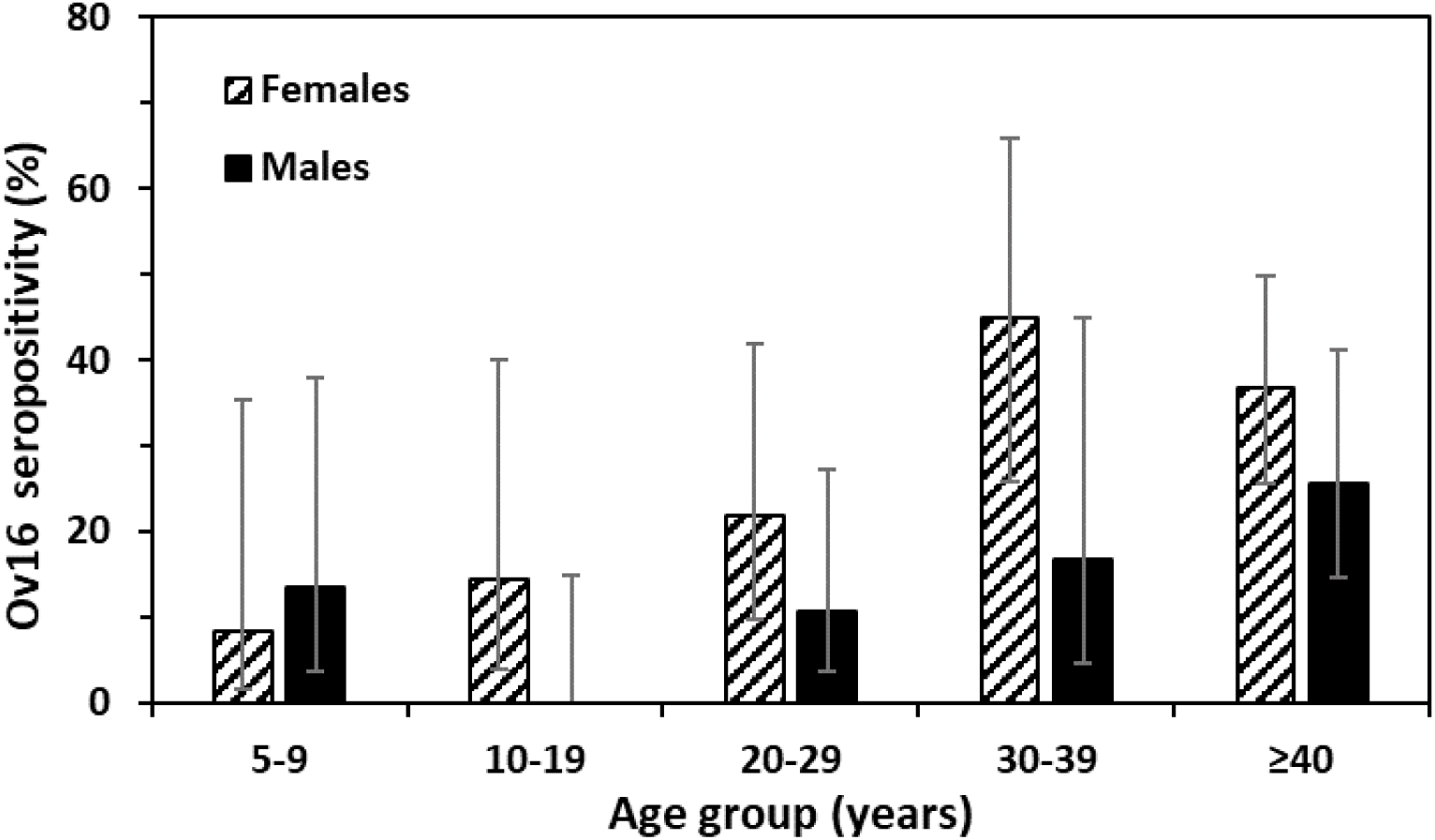
Ov16 seropositivity by age group and sex among 242 participants with recorded age. Hatched bars: Females, black bars: males, error bars: 95% confidence intervals.

**Table 3.** Age and sex distribution of test positivity for Ov16 RDT, skin-snip microscopy and nodule palpation for *Onchocerca volvulus* in the study communities, Bono Region, Ghana.

| Category | Positivity by Ov16 RDT<br>n +ve/N examined<br>(%) [95% CI] | Positivity by skin-snip<br>n +ve/N examined<br>(%) [95% CI] | Positivity by nodule palpation<br>n +ve/N examined<br>(%) [95% CI] |
| --- | --- | --- | --- |
| Age<sup>†</sup> (years) |  |  |  |
| 5–9 | 3/27 (11.1%) [3.9% – 28.1%] | 3/13 (23.1%) [8.2% – 50.3%] | 1/27 (3.7%) [0.7% – 18.3%] |
| 10–19 | 2/36 (5.6%) [1.5% – 18.1%] | 2/15 (13.3%) [3.7% – 37.9%] | 2/36 (5.6%) [1.5% – 18.1%] |
| 20–29 | 8/51 (15.7%) [8.2% – 28.0%] | 2/16 (12.5%) [3.5% – 36.0%] | 1/51 (2.0%) [0.4% – 10.3%] |
| 30–39 | 11/32 (34.4%) [20.4% – 51.7%] | 3/10 (30.0%) [10.8% – 60.3%] | 2/32 (6.3%) [1.7% – 20.2%] |
| ≥40 | 31/96 (32.3%) [23.8% – 42.2%] | 1/37 (2.7%) [0.5% – 13.8%] | 8/96 (8.3%) [4.3% – 15.6%] |
| Total | 55/242 (22.7%) [17.9% – 28.4%] | 11/91 (12.1%) [6.9% – 20.4%] | 14/242 (5.8%) [3.5% – 9.5%] |
| Sex<sup>‡</sup> |  |  |  |
| Female | 43/136 (31.6%) [24.4% – 39.9%] | 6/53 (11.3%) [5.3% – 22.6%] | 6/136 (4.4%) [2.0% – 9.3%] |
| Male | 17/117 (14.5%) [9.3% – 22.0%] | 5/41 (12.2%) [5.3% – 25.5%] | 8/117 (6.8%) [3.5% – 12.9%] |
| Total | 60/253 (23.7%) [18.9% – 29.3%] | 11/94 (11.7%) [6.7% – 19.8%] | 14/253 (5.5%) [3.3% – 9.1%] |

<sup>†</sup> Age was recorded for 242 of the 254 participants. <sup>‡</sup> Sex was recorded for 253 participants.

### Concordance between tests

Table 4 presents the results of the 2 x 2 concordance test between the Ov16 RDT assay and skin-snip microscopy for the 94 participants from whom skin biopsies were taken, as well as the results of Ov16 RDT compared with the ‘Oncho’ status of the same participants (combining the results of skin-snip microscopy and nodule palpation). In both cases, the value of kappa is not statistically different from zero, indicating poor agreement between tests after correcting for chance agreement [26]. The McNemar’s Chi-squared statistics and corresponding P-values indicate that the results of the tests are not significantly associated, with a proportion of positive agreement between 13% and 18%, a proportion of negative agreement of approximately 83%, and a proportion of overall agreement of roughly 70%. The sensitivity of the Ov16 RDT was about 20% and its specificity of about 80% in comparison to skin-snip microscopy or skin-snip and/or nodule palpation.

**Table 4.**
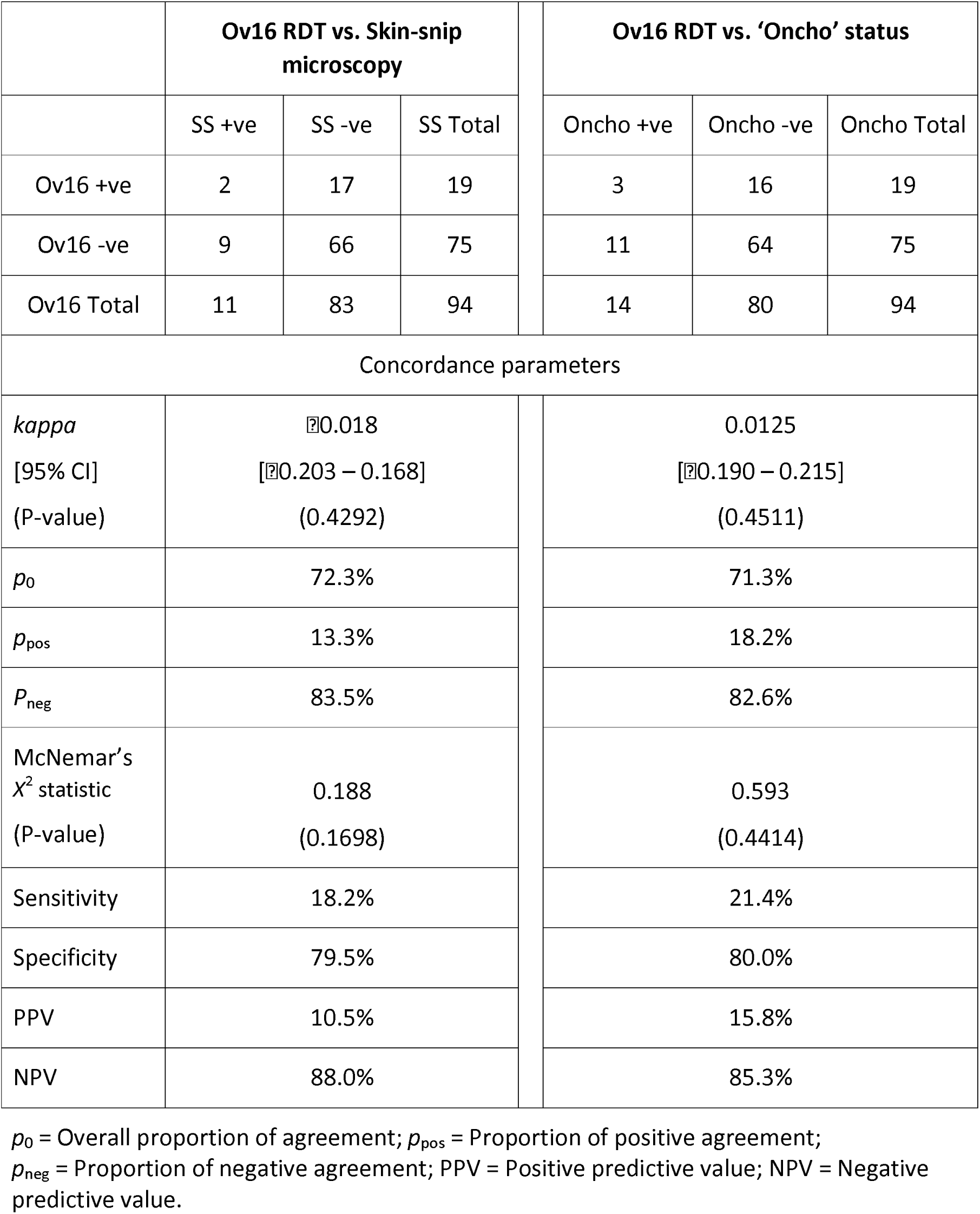
Two x two concordance results for the comparison between Ov16 Rapid Diagnostic Test (RDT) vs. skin-snip microscopy (SS), and Ov16 RDT vs. ‘Oncho’ status (skin-snip and/or nodule palpation).

### Acceptability of Ov16 RDT, skin-snip microscopy and nodule palpation

The 94 individuals who participated in all three tests were interviewed after the diagnostics were completed (exit questionnaire, S1 Text). The majority of the participants (71/94 = 75.5%) preferred the Ov16 RDT to skin-snip microscopy for reasons including that the Ov16 RDT procedure was less painful, less invasive and that they could see their results in 30–40 minutes, rather than waiting until the next field visit to learn whether or not they were ‘positive’:

> *“This test (Ov16 RDT) is better! It is less painful than the one where you (health personnel) ‘cut the buttocks’ (skin snip)” (Female participant, Abekwai 2).*
>
> *“I don’t normally see the need to participate in the test where you take some skin from my buttocks. A lot of times, you (health personnel) just cut the skin and we don’t hear anything about our results. I like this new one (Ov16 RDT) as I got to know my results immediately” (Male participant, Abekwai 3)*.

Despite the majority’s preference for Ov16 RDTs, 34% (32/94) remarked that they were ‘okay’ with skin-snip microscopy and were willing to ‘endure’ the pain, as long as they were convinced that undergoing the test would improve their health and wellbeing. This proportion of respondents was in agreement with the 37% of the recruited individuals who agreed to be skin-snipped (Table 1).

> “*For me, I am okay if you (technician) cut skin on my buttocks (skin snip), as long as it will help towards treating me early. The pain is only temporary, good health is what matters*” (Adult male, Attakrom).

However, 20% (19/94) of participants detested the skin-snip method and stated that they would not engage in any future surveys involving this method. The major reasons for this included the painful nature of skin-snipping and its invasiveness. Some participants also argued that they experienced itching, developed a rash or suffered from other allergic reactions after they had been skin-snipped:

> *“For me, I will run away if you (technician) come here again with that your scissors, it is too painful” (Female participant, Johnykrom)*.

Regarding the use of nodule palpation, 98% (92/94) of the study participants were ‘okay’ with it, with 18% (17/94) preferring to be examined for nodules by a person of the same sex, and 2% (2/94) not being at all comfortable with being palpated for nodules due to religious reasons:

> “*As for me, I don’t like it when the female doctor is touching parts of my body as she will see my nakedness” (Adult male, Blibor)*.

### Usability and acceptability of Ov16 RDT among technicians in the study

The study interviewed all nine technicians who performed the Ov16 RDTs and who were also experienced in skin-snip microscopy, with 89% (8/9) preferring the former test over the latter. The reasons for this included that it required less effort and training and met with less resistance from the survey participants on the account of being less painful and less invasive. The rapid turnaround time of the Ov16 RDT results (within 30–40 minutes from sampling to results) as opposed to more than 24 hours for skin-snip microscopy further increased the willingness of the community to take part in surveillance activities. The technicians also liked the fact that children were able to participate in the serological survey. Most of the technicians also indicated that they trusted the results from the Ov16 RDT as it tends to lend itself to fewer human errors. When prompted on the challenges with the use of the Ov16 RDT, most technicians referred to the fact that being antibody-based, the test could not differentiate between past and current infections. They therefore advocated for both tests; first Ov16 RDT for indication to exposure, followed by confirmation of active infection with skin-snip microscopy when necessary. Nevertheless, all technicians remarked that they would prefer to use Ov16 RDTs in future surveillance activities.

### Cost analysis for Ov16 RDT and skin-snip microscopy

#### Time considerations

The training of nine technicians for the Ov16 RDT took a maximum of two hours (120 minutes) in contrast to 14 days (6,720 minutes for an 8-hour daily schedule) for the training of five technicians on skin-snip microscopy. The Ov16 RDT took approximately 10 minutes to take a sample from each participant (labelling the cassette, selecting, and cleaning a finger, pricking the finger and placing the blood and the assay diluent in the cassette) and 30–40 minutes to get and communicate the results (20 minutes to read the results from the cassette and 10–20 minutes to record such results and convey them to participants). By contrast, skin-snip microscopy took about 30 minutes for sampling from each participant (cleaning left and right iliac crests with alcohol swabs, taking the skin snips with disinfected punches, placing the snips in labelled microtitre plate wells containing incubation medium, recording participant details), 24 hours for the incubation of the snips and about 20 minutes to pipette and observe the incubation medium under a microscope under two magnifications, for a total of 24 hours and 50 minutes (1,490 minutes) to obtain the results. A common concern was that the participants of skin-snip microscopy typically never get to know their results or at best get to know them only during the next field visit, which can take weeks or months. These have cost implications for both Ov16 RDT and skin-snip microscopy tests.

#### Cost considerations

The total cost for the activities targeting 400 participants was estimated at US$32,156. The cost was apportioned to Ov16 RDT, skin-snip microscopy and shared cost. The costs incurred for Ov16 RDT and skin-snip microscopy test and activities were US$2,412 (7.5%) and US$22,670 (70.5%), respectively. The shared cost incurred accounted for 22% of the total cost of the study (US$7,074). Within the total cost, 94.8% (US$30,484) corresponded to fieldwork cost, comprised of transport and lodging (72.0%), supplies, devices and instruments (21.4%), and labour cost (1.4%). Training and reporting costs accounted for 5.1% (US$1,640) and 0.1% (US$32) of the total cost, respectively. The total cost per participant of the Ov16 RDT was US$ 23.72 (US$6.03 for test-specific costs and US$17.69 for shared costs); the cost of skin-snip microscopy was US$74.36 (US$56.67 for test-specific costs and US$17.69 for shared costs). The cost per participant of deploying Ov16 RDT and skin-snip microscopy testing based on 400 participants and split by category is presented in Fig 3.

**Fig 3.**
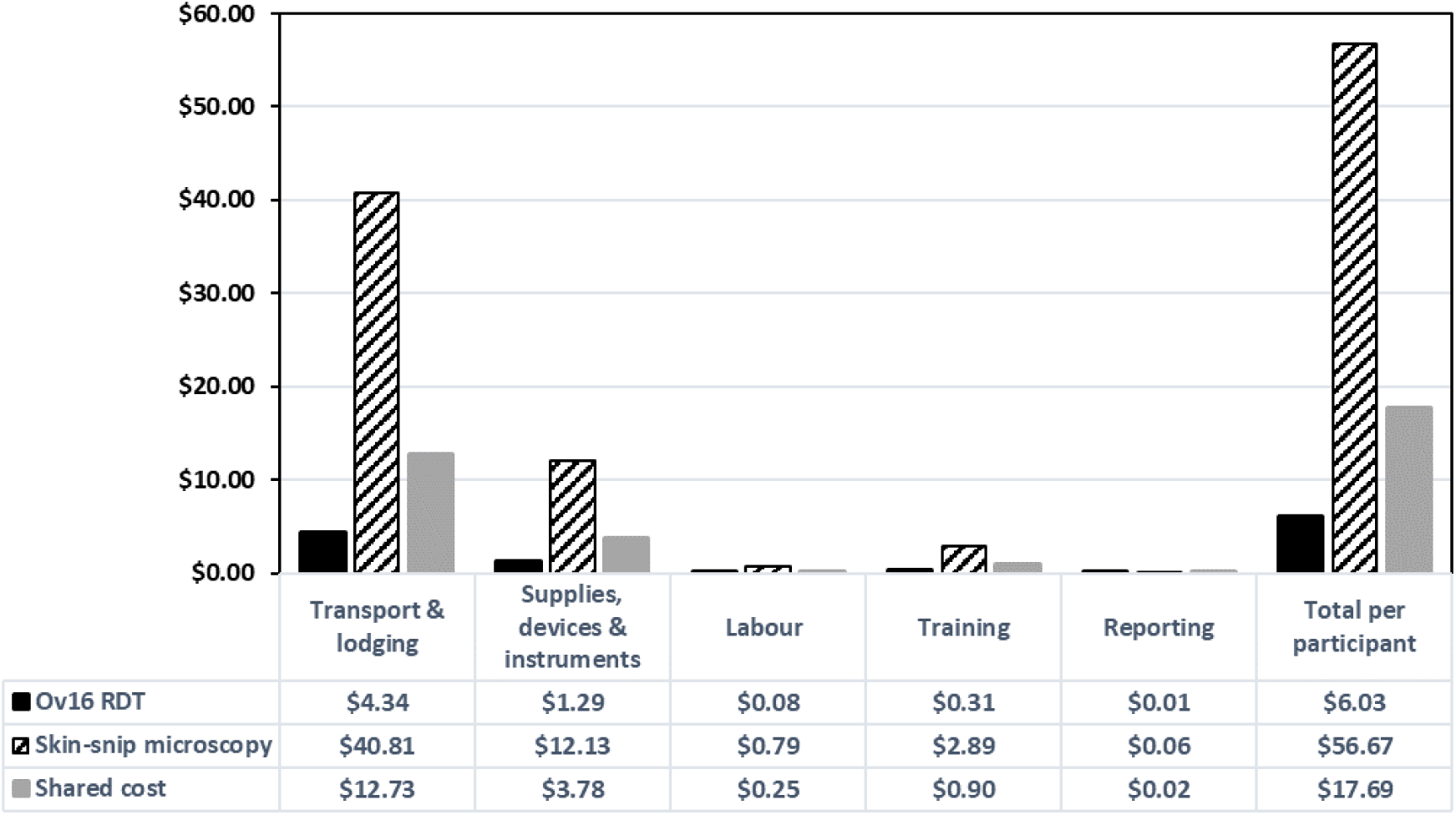
Cost analysis per Ov16 rapid test (Ov16 RDT) and skin-snip microscopy for 400 individuals tested for each diagnostic.

Proportionally reducing costs according to the numbers tested in our study (254 for Ov16 RDT and 94 for skin-snip microscopy) and adjusting for fixed inputs, the total cost of the activity was estimated at US$24,708 for the 6 communities surveyed, with a net cost per participant of US$27 for Ov16 RDT and US$207 for skin-snip microscopy, highlighting the impact of the volume effect [10].

## Discussion

This study aimed to determine the usability, acceptability and cost of the Ov16 RDT for onchocerciasis surveillance in endemic communities in the middle belt of Ghana, particularly in the Tain District and Wenchi Municipality where it has been reported that onchocerciasis persists after 27 years of ivermectin MDA [19,20]. The seroprevalence among study participants was 24%, the microfilarial prevalence was 12% and the nodule prevalence was 6%. Sex, age group, occupation and palpable nodule status were significantly associated with Ov16 status, whereas duration of stay in the village, living in proximity to a stream, visiting the stream, frequency of visiting the stream and skin-snip status were not (Table 2). Among the Ov16 seropositives, there was a significantly higher proportion of females, those aged 30 years or older, living near streams, visiting such streams and engaged in farming. However, the proportions of those presenting skin microfilariae or palpable nodules were significantly lower.

Since the study communities have received ivermectin MDA for nearly three decades, those aged ≥30 years at the time of our study would have been exposed as children to baseline levels of transmission. We have previously reported that the study area was initially mesoendemic, with a baseline microfilarial prevalence of 41% to 48% [20]. This is broadly in agreement with the 33% seroprevalence we found among those 30 years or older. Of the 126 females sampled, more than half (61%) were aged ≥30 years, whereas of the 116 males sampled, less than half (44%) belonged to this age group; nearly 100% of those 30 years of age or older were farmers (76/77 females and 50/51 males).

As the blackfly vectors breed in fast-flowing rivers and streams [1], it is perhaps not surprising that those living near or visiting such water bodies were more frequently represented among seropositive individuals, but contrary to expectation the proportion of those visiting streams daily was lower than that of those visiting less often. Farming can be another activity that increases exposure to blackfly bites, and historically, agricultural communities are known to have been highly endemic for onchocerciasis as farms are often situated near rivers that provide water for crops [29]. As blackflies are outdoor and diurnal biters, farmers working outdoors for several hours of the day may have an increased risk of exposure. In the Tombel Health District, Southwest region of Cameroon, and after 15 years of CDTI, Nyagang et al. [30] reported higher onchocerciasis prevalence in those aged more than 60 years, who were mostly farmers. The greatest prevalence of infection (2.1%) was among farmers [30]. A study in Southwest Ethiopia, also after 15 years of CDTI, reported that individuals aged ≥35 years had a microfilarial prevalence of 15.7% compared to 1.4% in those aged 15–24-years (no children under 15 were examined), and that approximately 60% of the participants were farmers, of whom 9% were microfilaria-positive [31]. The authors stated that adults were engaged in outdoor activities, increasing their exposure to blackfly bites [31].

Although Ov16 seroprevalence levels, particularly in the young, have been considered to indicate exposure to infection, knowledge of the parasite stage(s) that elicit IgG4 antibody seroconversion to the Ov16 antigen, as well as of the dynamics of seroconversion and potential seroreversion or antibody decay under transmission interruption remains outstanding. Willen et al. [32,33] reported, using data from Ghana, on the development of an IgG-based immunoassay to quantify antibodies to *Simulium damnosum* sensu lato saliva that, if combined with assays to detect past or current exposure to *O. volvulus* could help to investigate exposure to blackfly bites and to the parasite.

In our study, Ov16, skin-snip and nodule positivity were generally higher among those aged 30 years or older (with the exception of microfilarial prevalence, which was lowest in those ≥40 years of age). Since only 94/254 (37%) of the participants agreed to be skin-snipped, with complete data on age and sex for 91 of these, our results need to be interpreted with caution. Notwithstanding our small sample sizes, 11%, 23% and 4% of the 5–9-year olds examined were positive for antibodies against Ov16, microfilaridermia and onchocercomas, respectively (Table 3). As the nodule-positive child was aged 5 and the pre-patent period (from infection to detectable microfilaridermia) of *O. volvulus* has been estimated to range between 1 and 3 years [34], we can infer that this child became infected when he was 3–4 years of age, underscoring the occurrence of ongoing transmission in the study villages. Although Ov16 seroprevalence by age was generally higher for females compared to males (with the exception of the 5–9-year age group), sample sizes were small, leading to wide 95%CIs (Fig 2), precluding us from drawing firm conclusions about sex- and age-specific exposure patterns.

The low positive agreement between Ov16 and skin-snip positivity is likely explained by the fact that treatment leads to a reduction in skin microfilariae but antibodies to Ov16 may remain for much longer [10], with the levels of Ov16 and skin-snip positivity becoming increasingly disassociated as treatment progresses.

The McNemar chi-square statistic showed no association between Ov16 RDT and skin-snip microscopy (or ‘Oncho’ status when combining skin-snip microscopy and/or nodule palpation), indicating that Ov16 RDT is not an appropriate tool for determination of active infection trends in surveillance activities, for which, in any case, it was not designed. However, given the relatively high level of negative agreement (83%–84%) and NPV (85%– 88%), it may be possible to use Ov16 RDT for screening (all ages), subsequently focusing on those Ov16 positive for further testing with skin-snip microscopy and/or skin-snip PCR. According to the RDT manufacturer, the test has 81% sensitivity and 99% specificity [10], but likely due to long-term treatment in the study area, our estimated values were very low (about 20% and 80%, respectively) (Table 4).

We acknowledge that the WHO guidelines for stopping MDA and verifying elimination of transmission do not advocate the use of Ov16 serology to detect active infection or its use in all ages, but rather in those under 10 years of age using ELISA, with a seropositivity <0.1% at the upper 95% confidence bound in a sample of 2,000 children being taken as an indication of transmission interruption [35]. Modelling studies have investigated the applicability, to predict EOT outcomes, of this (and less stringent) threshold(s) and age group(s) under different assumptions of exposure and regulation of parasite abundance [36], and ongoing studies (some in Ghana) are being conducted to test such thresholds in the field.

We found that conducting Ov16 RDT testing in all ages provided useful information interpretable in the context of the duration of treatment in the area, as done by others in Mali and Tanzania [37,38]. Our results also indicate that the study communities are not yet at the point of commencing Stop-MDA surveys despite having received nearly three decades of ivermectin MDA. However, since we did not sample 2,000 children aged less than 10 years, our results are surrounded by considerable uncertainty. Despite this uncertainty, we have documented persistence of *O. volvulus* infection and associated clinical manifestations in the study area [19,20], and investigated factors influencing treatment adherence which could partly explain such persistence [21]. Entomological studies should also be undertaken in the study area to assess infection levels in simuliid population samples.

Studies done in other African settings with long-term CDTI (using the Ov16 RDT with whole blood) have reported, in the Ulanga district of Morogoro, Tanzania (an initially hyperendemic area), seroprevalence levels of 14% (27/191) in those aged 6–10 years and 33% (26/79) in those aged 11–12 after more than 20 years of CDTI [39]. In a formerly highly endemic area in the Mbam and Sanaga river valleys of Cameroon, Siewe Fodjo et al. [40] found an Ov16 seropositivity of 47% (68/145) in Bilomo (Centre Region) and 52% (13/25) in Kelleng (Littoral Region) among children aged 7–10 years after >13 years of CDTI. A factor likely involved in the persistence of onchocerciasis in this area is the very high (and perennial) biting rates of the blackfly vectors, as documented in other localities of the Mbam and Sanaga river systems, both historically and recently [41,42]. As our study area was originally mesoendemic, and annual biting rates in the region were of the order of 3,000–4,000 per person [19], it would be advisable to update entomological studies in the Tain District and Wenchi Municipality of Ghana to better understand the potential contribution of blackfly abundance, species composition and transmission intensity to persistent infection. A study in the Nkoranza North District (see Fig 1) documented highly seasonal transmission, with monthly biting rates of approximately 800 in the peak transmission season and 0–100 in other months of the year [43].

In terms of diagnostic test acceptability, most participants preferred the Ov16 RDT to skin-snip microscopy and nodule palpation on account of its being less invasive, less painful and more rapidly generating results that the participants could see *in situ*. These attributes of the RDT could be capitalised upon to increase the willingness of the population to participate in surveillance activities. Our findings are consistent with those of Dieye et al. [10] in south-eastern Senegal, who reported a greater willingness of participants to take part in onchocerciasis surveillance activities involving Ov16 RDT. Interestingly, the participation rates in Ov16 RDT (99.7%) and skin-snip microscopy (32.7%) in the Senegal study [10] are very similar to ours in Ghana (100% and 37%, respectively), suggesting that about a third of the population might be willing to participate in skin-snipping surveys. As endemic countries transit the path towards onchocerciasis EOT, it is increasingly important that the surveillance tools used in control and elimination programmes are both effective and acceptable. Residents of meso- and hyperendemic communities have typically had a long history of MDA treatment and are, therefore, increasingly apathetic towards control and surveillance activities, especially because the disease is no longer perceived as a major public health problem in some areas.

As mentioned, about a third of the individuals taking part in the ‘exit’ interviews remarked that although skin-snipping is painful, they would be willing to participate in future parasitological surveys using skin-snip microscopy. This was partly due to the research team members explaining to the participants the need for the test and the potential benefits to their health outcomes. Thus, the influence of the surveillance staff on participation rates should not be underestimated. The National Onchocerciasis Control Programme could leverage this finding to tackle issues of growing apathy in endemic communities.

The Ov16 RDT was also generally preferred among the technicians involved owing to its being relatively simple and easy to use, quick turnaround and potential for increasing community participation in future surveys, in agreement with the study in Senegal [10]. However, the technicians in our study were well aware of the Ov16 RDT limitations concerning its inability to distinguish between past and present exposure/infection.

For a total of 400 individuals, the total cost per participant was estimated as US$24 for Ov16 RDT and US$74 for skin-snip microscopy, the former being approximately a third of the cost of the latter. The difference in cost between the two tests per individual (US$50) was mainly driven by differences in transport and lodging, supplies, devices and instruments, labour and training. Regarding training requirements, Ov16 required a relatively short time (approximated 2 hours) to train nine technicians. By contrast, skin-snip microscopy training (sample-taking technique and microscopy) took 14 days. Also, supplies, devices and instruments needed for skin-snip microscopy include expensive instruments such as corneoscleral punches, microscopes, microtitre plates, pipettes etc., whereas those for Ov16 RDT principally include the kits from the manufacturer. Although the cost of transport and lodging was considered jointly for the deployment of the two tests and allocated according to the proportion of costs for each specific test and their shared costs, it is important to note that if Ov16 RDT alone were used in typical surveillance, fewer days would likely be necessary, making the Ov16 RDT even less costly. Nevertheless, challenges with Ov16 RDT procurement must be considered when planning surveillance activities.

### Limitations

Our study has a number of limitations that should be addressed in future work. Firstly, study participants were not randomly sampled and, therefore, study results need to be interpreted with caution as they may not be truly representative of the study communities. Sample sizes to ensure a given power or precision were not calculated before the study; only people who agreed to gather at the focal point in their communities were included in the study (convenience sampling), with only 12% of the population participating overall (Table 1). This likely led to bias. Participants with clinical manifestations seeking to be examined by the research team may also be those with greater levels of infection, leading to overestimation of prevalence levels. Alternatively, individuals who have greater adherence to treatment may also have greater willingness to participate in surveillance activities, with the surveys missing a proportion of those who have higher levels of infection, resulting in underestimation. Secondly, social desirability bias may have influenced the results of the usability and acceptability surveys, as they were based purely on verbal responses. Thirdly, the slides prepared from the incubation medium of the skin snips for detection of *O. volvulus* microfilariae were not fixed and stained to assess their characteristic morphological features, and no skin-snip PCR was conducted to confirm species-specific identification. Therefore, we cannot totally rule out the presence of other filarial species in the skin snips such as *Mansonella streptocerca* (skin-dwelling microfilariae), or even *M. perstans* (blood-dwelling microfilariae that may be present in the event that snips were not wholly bloodless). Fourthly, we used the Ov16 RDT with whole blood, but collecting dried blood samples that are eluted in the lab and used in the RDT may reach a sensitivity similar to that of the Ov16 ELISA recommended by the WHO [44]. However, this procedure would increase the reading time of the results to 30 minutes–24 hours, with the latter recommended for increased accuracy of seroprevalence estimates [44,45]. Finally, as our cost estimates were calculated using specific costs for Ghana, there is a need for similar implementation research in other endemic countries and settings, as exemplified by the very different cost estimates presented for Senegal [10].

## Conclusions

This study has demonstrated the usability of the Ov16 RDT to identify ongoing transmission using seroprevalence in children aged 5–9 years in an area with nearly three decades of CDTI, and more generally, exposure patterns in the general population. The test was also more acceptable and feasible than skin-snip microscopy, less costly to implement, and may help overcome the increasing reluctance of endemic communities to participate in onchocerciasis surveillance activities. Regardless of these advantageous features, further R&D efforts are necessary to increase the diagnostic performance of serological tests and improve their ability to distinguish between past and patent infection [46]. Ideally, such tests could be used to screen the population and focus on those seropositive for further parasitological testing, or if it can be ensured that the measured antibody responses are elicited by microfilariae and mirror their dynamics following treatment, to obviate this need altogether.

## Data Availability

All the data are contained in the Tables, Figures, and Supporting information.

## Abbreviations

CDTI: Community Directed Treatment with Ivermectin
CI: Confidence interval
DALY: Disability Adjusted Life Year
ELISA: Enzyme-Linked Immunosorbent Assay
EOT: Elimination of Transmission
GBD: Global Burden of Disease
IgG: Immunoglobulin G
IgG4: Immunoglobulin G4
MDA: Mass Drug Administration
NPV: Negative predictive value
NTD: Neglected Tropical Disease
OCP: Onchocerciasis Control Programme in West Africa
PCR: Polymerase Chain Reaction
PES: Post-Elimination Surveillance
PPV: Positive predictive value
PTS: Post-Treatment Surveillance
R&D: Research and Development
REMO: Rapid Epidemiological Mapping of Onchocerciasis
RDT: Rapid Diagnostic Test
SSA: Sub-Saharan Africa
UI: Uncertainty interval
WHO: World Health Organization

## Supporting information

**S1 Text.** Interviewer guide for participants’ and technicians’ ‘exit’ interviews after conducting tests.

## Authors’ contributions

**Conceptualization:** Kenneth Bentum Otabil, María-Gloria Basáñez, Henk D.F.H. Schallig, Robert Colebunders.

**Data curation:** Kenneth Bentum Otabil, María-Gloria Basáñez.

**Formal analysis:** Kenneth Bentum Otabil, María-Gloria Basáñez, Lydia Datsa, Andrews Agyapong Boakye, Robert Colebunders.

**Funding acquisition:** Kenneth Bentum Otabil, María-Gloria Basáñez, Henk D.F.H. Schallig Robert Colebunders.

**Investigation:** Kenneth Bentum Otabil, Ameyaa Elizabeth, Michael Oppong, Prince Mensah, Richmond Gyasi-Ampofo, Emmanuel John Bart-Plange, Theophilus Nti Babae, Michael Tawiah Yeboah, Prince Nyarko, Prince Charles Kudzordzi, Anabel Acheampong, Edwina Twum Blay.

**Methodology:** Kenneth Bentum Otabil, María-Gloria Basáñez, Henk D.F.H. Schallig, Robert Colebunders.

**Project administration:** Kenneth Bentum Otabil.

**Resources:** Kenneth Bentum Otabil, María-Gloria Basáñez, Henk D.F.H. Schallig, Robert Colebunders.

**Supervision:** María-Gloria Basáñez, Henk D.F.H. Schallig, Robert Colebunders.

**Visualization:** Kenneth Bentum Otabil, María-Gloria Basáñez, Lydia Datsa, Andrews Agyapong Boakye.

**Writing – original draft:** Kenneth Bentum Otabil, Maria-Gloria Basáñez.

**Writing – review & editing:** Kenneth Bentum Otabil, María-Gloria Basáñez, Ameyaa Elizabeth, Michael Oppong, Prince Mensah, Richmond Gyasi-Ampofo, Emmanuel John Bart-Plange, Theophilus Nti Babae, Lydia Datsa, Andrews Agyapong Boakye, Michael Tawiah Yeboah, Prince Nyarko, Prince Charles Kudzordzi, Anabel Acheampong, Edwina Twum Blay, Henk D.F.H. Schallig, Robert Colebunders.

## Acknowledgements

We thank all the staff and students of the Department of Biological Science and the Centre for Research in Applied Biology who provided support in one way or another to ensure the successful implementation of this research. We also appreciate the efforts of the directors and staff of the Wenchi Municipal Health Directorate, Nsawkaw District and the Subinso Health Centre for their enormous contributions towards the study. Our appreciation also goes to the chiefs and people of the study communities who partnered with us in this study.

## Funding

This project was supported by a small grant from the Royal Society of Tropical Medicine and Hygiene, UK awarded to KBO in 2020, and from the United States Agency for International Development (USAID) and UK FCDO from the British people (UK FCDO) through the Coalition for Operational Research on Neglected Tropical Diseases (COR-NTD) and administered by the African Research Network for Neglected Tropical Diseases (ARNTD). MGB acknowledges funding from the MRC Centre for Global Infectious Disease Analysis (MRC GIDA, grant number MR/X020258/1), funded by the UK Medical Research Council (MRC). This UK-funded award is carried out in the frame of the Global Health EDCTP3 Joint Undertaking. RC received funding from Research Foundation Flanders (FWO, grant number G0A0522N).

## Competing interests

The authors declare that there are no competing interests regarding the publication of this article.

## Ethical approval

This study was approved by the Committee for Human Research and Ethics at the University of Energy and Natural Resources, Sunyani, Ghana (approval number: CHRE/AP/012/021). Written informed consent was obtained from all participants before recruitment. All procedures in this study were in accordance with the ethical standards of the Helsinki Declaration (1964, amended most recently in 2008) of the World Medical Association.

## Availability of data and materials

All the data are contained in the Tables, Figures, and Supporting information.

## Notes

### Competing Interest Statement

The authors have declared no competing interest.

### Author Declarations

The Committee for Human Research and Ethics at the University of Energy and Natural Resources, Sunyani, Ghana gave ethical approval for this work (approval number: CHRE/AP/012/021).

