## Supplementary Text 1 for "Usability, acceptability and cost of the SD BIOLINE Ov16 rapid diagnostic test for onchocerciasis surveillance in endemic communities in the middle belt of Ghana"

**S1 Text. Interviewer guide for participants' and technicians' interviews**

**Objective:** To gather data on user (participant/technician) experience in relation to the Ov16 RDT, how the test was received by participants/technicians, and how it compared with skin-snip microscopy and nodule palpation.

**Method:** Interviews with community members and technicians were recorded as audio files, transcribed and translated from local language (*Twi*) into English. Interview data were coded using content analysis based on key themes from the semi-structured interviews.

**Guide questions:**

1. Did you take part in Ov16 RDT, skin-snipping and nodule palpation in this study?
2. Which of these do you think is the best? Explain why.
3. What aspects you liked and/or disliked about each test?
4. Will you participate in future onchocerciasis surveillance activities that use Ov16 RDT, skin-snipping, and/or nodule palpation? Explain your answer.
5. What influences whether you participate in the test or not?
6. Is there anything else you would like to say about each of the tests?
